# Towards a rapid-turnaround low-depth unbiased metagenomics sequencing workflow on the Illumina platforms

**DOI:** 10.1101/2023.01.02.22283504

**Authors:** Winston Koh Lian Chye, Si En Poh, Chun Kiat Lee, Tim Hon Man Chan, Gabriel Yan, Kiat Whye Kong, Lalita Lau, Wai Yip Thomas Lee, Clark Cheng, Shawn Hoon, Yiqi Seow

## Abstract

Unbiased metagenomic sequencing is conceptually well-suited for first-line infectious disease diagnostics because it does not require guesswork on the causative organism and can cover all known and unknown infectious entities other than prions. However, costs, turnaround time and human background reads in complex biofluids such as plasma stand in the way of more widespread deployment. Many protocols also require separate library preparations for DNA and RNA analytes, increasing the costs for detection. In this study, we developed a rapid unbiased metagenomics next-generation sequencing (mNGS) workflow that addresses these issues with a human background depletion kit (HostEL) and a library preparation kit that processes DNA and RNA simultaneously (AmpRE). In our analytical validation of the HostEL workflow, we were able to enrich and detect the signal from bacteria and fungi standards spiked in at physiological levels in plasma with low-depth sequencing (< 1 million reads). Our clinical validation also shows that 93% of plasma samples agreed with the clinical diagnostic test results when the diagnostic qPCR had a Ct < 33. The effect of different sequencing times was evaluated with the 19-hour iSeq 100 paired end run, a more clinically palatable simulated iSeq 100 truncated run and the rapid 7-hour MiniSeq platform. Significantly, our results demonstrate the ability to detect both DNA and RNA pathogens with low-depth sequencing. In conclusion, we demonstrate that iSeq 100 and MiniSeq platforms are compatible with unbiased low-depth metagenomics identification with the HostEL and AmpRE workflow and can be chosen based on required turnaround times.

## Introduction

Infectious disease is a worldwide health burden, with just the eight major infectious diseases (HIV, malaria, measles, hepatitis, dengue fever, rabies, tuberculosis and yellow fever) causing more than 156 million life-years lost in 2016 (1). Thus, early detection and diagnosis is important for timely interventions and treatment. Conventional methods, such as culturing of isolated pathogens and molecular amplification of nucleic acids, are widely used in diagnosis, but these methods require prior knowledge or suspicion of the pathogens (2–5). In contrast to the traditional methods, unbiased metagenomic next-generation sequencing (mNGS), as opposed with a targeted sequencing approach (6), can not only detect multiple pathogens in a single assay, but also discover novel or unexpected pathogens, like the recent outbreak of COVID-19 (3,7–19). Conceptually, a dependable robust metagenomic sequencing solution is an ideal first-line diagnostic if costs are low, time-to-result is fast and sensitivity is high.

Although genome sequencing technologies are developing rapidly (20–26), there are certain limitations involved. First, the approaches normally take a few days to complete, due to the complex processes of host background depletion, library preparation and data analysis (3,4). Second, as the samples contain high human background, deep sequencing needs to be done to increase sensitivity, which contributes to the high cost incurred (5,27,28). Although there are partial solutions available on human background depletion, strategies only focus on enriching bacteria DNA (28,29), which limits the utility when DNA or RNA viruses are causative. Third, most library preparation kits either uses DNA or RNA as input, but viruses can be both RNA and DNA-based, thus the need to deploy separate workflows for DNA and RNA preparation adds manpower time and costs to comprehensive unbiased sequencing.

To address these, we developed a workflow consisting of a HostEL (<u>Host El</u>imination) kit – a human background depletion strategy that also allows for enrichment of viruses as well; and <u>Amp</u>lification-<u>R</u>estriction <u>E</u>ndonuclease fragmentation (AmpRE) – a single tube DNA/RNA library preparation. HostEL uses magnetic bead-immobilized nucleases to deplete human background after selective lysis, replacing enzyme deactivation with magnetic pull-down, to enrich both pathogen DNA and RNA. AmpRE is a pure additive process for DNA and RNA input with only two clean-up steps before the library is ready for sequencing. It can reduce the processing time, while maintaining pathogen signal when performing shallow sequencing on iSeq 100, which allows more rapid diagnosis of communicable diseases. With the combination of the two strategies, our method not only can detect bacteria, but also fungi, DNA and RNA viruses.

We conducted an analytical validation for the workflow using plasma with artificial spiked-ins (ZymoBIOMICS Microbial Community Standard) at various concentration to mimic the physiological infection conditions down to 70 copies/μL of plasma (30). The samples were sequenced on two sequencing platforms, iSeq 100 and MiniSeq, with 3 different sequencing turnaround time to cater to different clinical workflow needs and demonstrated high sensitivity and specificity with each platform. We also conducted a clinical validation with 42 plasma samples, with 93% of the samples agreeing with the qPCR results on the clinical samples.

## Materials and Methods

### Analytical validation

ZymoBIOMCS Microbial Community Standard (Supplementary Table 1), consisting of 8 bacterial and 2 fungal strains at 1.4 × 10^7^ cells/μL, was diluted 100x (1.4 × 10^5^ cells/μL), 250x (5.6 × 10^4^ cells/μL), 1000x (1.4 × 10^4^ cells/μL) and 2000x (7.0 × 10^3^ cells/μL) before spiking 5μL into 500μL of human plasma. These gave a microbial spike in of concentrations of 1,400,000 cells, 560,000 cells, 140,000 cells, and 70,000 cells per mL of human plasma respectively. The spiked-in plasma was either HostEL-depleted (Aptorum Innovation) or just bead beaten. For HostEL-treated samples, 30μl of incubation buffer and 10μl of nuclease beads were mixed with 500μl of plasma amd incubated for 20 minutes shaking at 37°C. The nuclease beads were then with a magnetic rack and supernatant bead-beaten for 30s before extraction using Zymo Quick DNA/RNA Viral extraction kit (Zymo Research), eluting in 30μl. The control samples were just bead-beaten before extraction. Libraries were prepared with AmpRE kit (Aptorum Innovations) as per manufacturer’s instructions. Five and twenty samples were pooled and sequenced on iSeq 100 and MiniSeq respectively. Using 7μl of TNA as input, first strand synthesis in 10μl reaction is followed by the addition of 10μl 2^nd^ strand reagent for second strand synthesis, then an addition of 30μl amplification mix was added for amplification with methylated nucleotides. Cleanup with SPRI beads and elution in 10μl was followed by digestion for 4μl of eluted DNA in a 5μl reaction for restriction digest of methylated nucleotides. 10μl of ligation buffer was added to the fragmented DNA for ligation and tailing of sequencing arms before amplification and barcoding. The sample was then cleaned up using SPRI beads and eluted in 10μl and quantified on Agilent 4150 TapeStation Instrument (Agilent Technologies) using Agilent D5000 ScreenTape System (Agilent Technologies).

### Sequencing on iSeq 100 System and MiniSeq System

Each quantified sample was diluted to a concentration of 190pM with 10 mM TrisHCl, pH 8.5. The samples were then pooled together in equal volume, making up to a total of at least 20μL. 20μL of the pooled sample was loaded onto the iSeq 100 cartridge before loading onto iSeq 100 for sequencing (31). For MiniSeq, each quantified sample was diluted to a concentration of 10nM with RSB (10mM Tris-HCl, pH 8.5 with 0.1% Tween 20) and pooled at equal volume, before diluting down to 1nM with RSB. The pooled library was denatured by mixing 5μL of 1nM pooled sample and 5μL 0.1N NaOH, and incubating at room temperature for 5 minutes. 5μl of 200mM Tris-HCl, pH 7.0 was then added to the denatured library. The denatured library was diluted down to 5pM by adding 985μL of pre-chilled Hybridization buffer (HT1) to the 15μL of denatured library. The library was further diluted down to loading concentration of 1.6pM by mixing 160μL of 5pM denatured library with 340μL of prechilled HT1. 500μL of the diluted sample was loaded onto the MiniSeq cartridge before putting onto the system for sequencing (32).

### Data Analysis

Fastq sequence files were exported from the sequencers and quality control was performed using Prinseq tool (33). The high quality data was obtained by removing the low complexity reads and then aligned to the human genome using HiSAT2 (34) Sequences that matched the human genome were removed, leaving behind the microbial sequences. The pathogens were then identified using KRAKEN2 and BRACKEN tools, aligning with NCBI reference genomes, and visualized using R script. For the simulated iSeq 100 run, the reads were trimmed to 100-base pair-end reads before processing through the same pipeline.

### Sample collection and Ethics statement

The studies involving human participants were reviewed and approved by NHG DSRB Study Reference Number: 2017/00632. The patients/participants provided their written informed consent to participate in this study. 5mL of blood plasma was collected in EDTA tubes. 500μL was aliquoted out and stored at -80°C before HostEL and AmpRE (Aptorum Innovations) processing as per manufacturer’s instructions. The samples were prepared as for analytical validation, but extracted with EZ1 Virus Mini 2.0 kit (Qiagen) instead, eluting in 60μl of water.

### qPCR validation

1μL of extracted samples and 0.3μM of each primer for pathogen (Supplementary Table 3) was added to each 20μl reaction. For bacteria, fungi and DNA viruses, Maxima SYBR Green / ROX qPCR Master Mix (Thermoscientific) was used. For RNA virus, Toyobo Thunderbird One Step qRT-PCR kit (Toyobo) was used with 1x SyBr Green (Thermoscientific). The mixture was amplified on the QuantStudio 1 qPCR system using the thermal profile: 50°C for 15 minutes, 95°C for 2 min for the initial denaturation, followed by 60 cycles of 95°C denaturation (15 s) and 60°C annealing and extension (60 s). Melting curve analysis was performed by ramping the temperature up to 95°C after qPCR to verify the specificity and identity of each of the PCR product. The list of primers used is shown in Supplementary Table 3.

## Results

### Low depth sequencing can detect physiological levels of bacteria

The mNGS workflow we used includes an hour-long host depletion stage that utilizes nuclease beads and selective lysis (Figure 1a) before bead-beating and total nucleic acid (TNA) extraction. Sequencing libraries are then generated with a 4-hour additive one-pot additive library preparation before sequencing in batches of 5 samples on iSeq 100, a low depth sequencing platform which can produce ∼4 million reads in 19h, to achieve low depth <1 million reads per sample. The iSeq 100 was selected for the proof-of-principle as it utilizes a plug-and-play cartridge that is user-friendly for the clinical laboratory. The reads were aligned to the human genome to discard background before quantifying and identifying pathogen signal in under 2 hours (Figure 1b).

**Figure 1.**
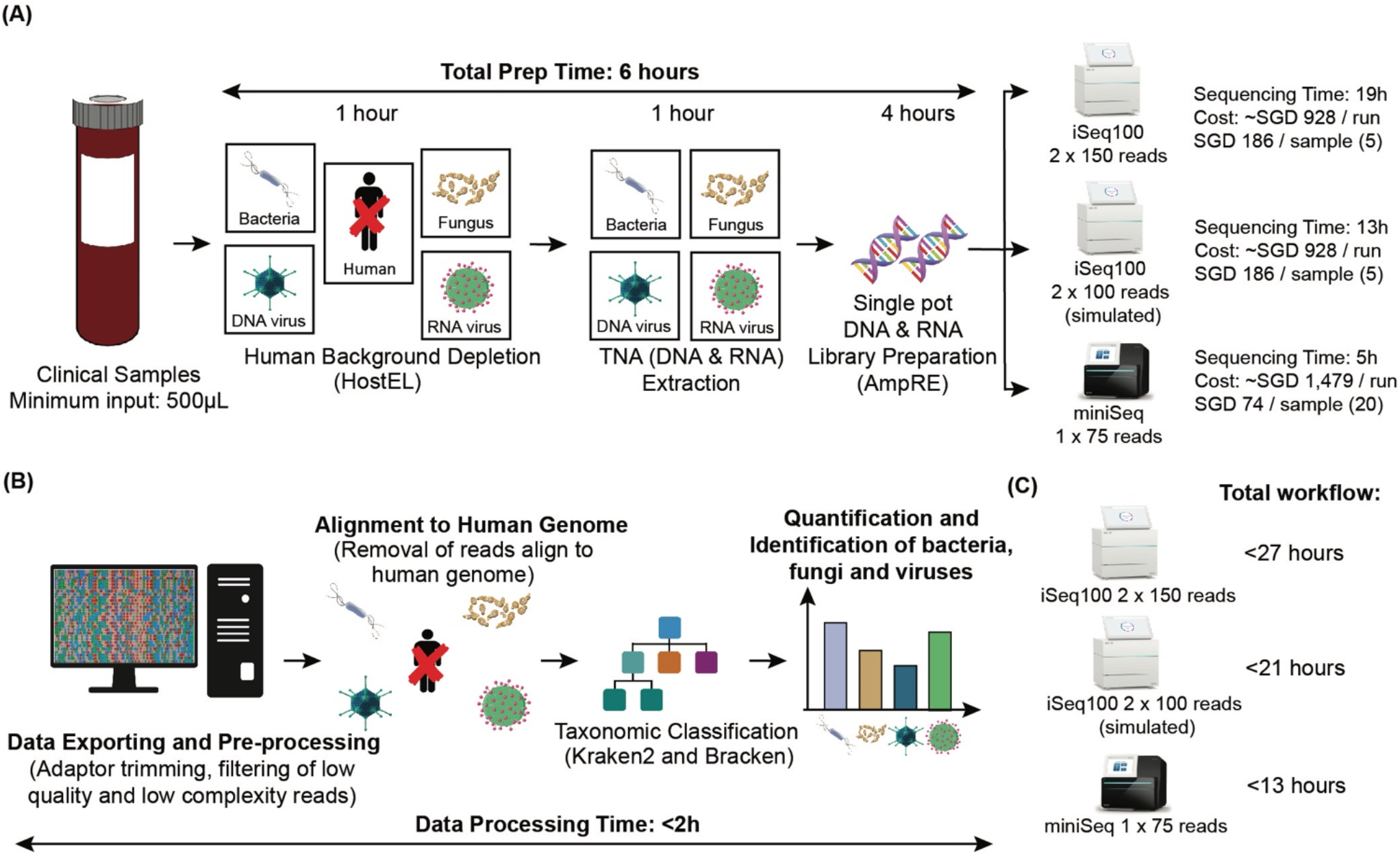
Schematic diagram of mNGS workflow using HostEL and AmpRE. (A) Workflow of mNGS using HostEL and AmpRE. A minimum of 500μl of a clinical sample, such as plasma is treated with HostEL to deplete human background before the total nucleic acids is extracted. TNA is then amplified into a sequencing library in a one pot AmpRE reaction, followed by sequencing on the iSeq 100 (2 × 150; 2 × 100 simulated) or MiniSeq (1×100). The time required for each step and the cost of sequencing are indicated. (B) Workflow of data analysis. Sequenced data is exported in Fastq format from the sequencing platforms, and high quality and compatibility reads are selected. These sequences are then aligned to human genome, where the matched human sequences are removed. The reads of the pathogens (bacteria, fungus, and both DNA and RNA viruses) are then quantified and identified using KRAKEN2 and BRACKEN tools. The whole process takes less than 2h to complete. (C) Comparison of the different sequencing platforms on the total workflow time.

To validate our workflow (Figure 2a), human plasma were spiked with various dilutions (100x, 250x, 1000x and 2000x) of ZymoBIOMICS Microbial Community Standard, which comprises of bacteria and fungi (Supplementary Table 1, Supplementary Figure 1), to achieve the total cell counts of 1.4 million, 560,000, 140,000, and 70,000 cells per mL of plasma respectively. This mimics the physiological concentrations of 10^5^ copies/mL in the body during active infection (30). Spiked-in samples without HostEL treatment, and sample with no spike-in were prepared as negative controls for comparison, which made up a total of 10 samples. Even with a median of 56498 paired end reads (2 × 150bp) per sample across all samples on the iSeq 100, the pipeline identified all 10 species at all dilutions when treated with HostEL while preserving relative distribution of species. In contrast, the untreated samples started losing signal at 140,000 and 70,000 cells per ml (Figure 2b). The enrichment is ∼2-fold at 1.4 million cells/mL of plasma of HostEL treated samples and extent to ∼4-fold at the lower concentrations on iSeq 100, while the enrichment on MiniSeq is at least 1.5-fold (Supplementary Table 2). In this study, the lowest limit of detection (LOD) was set to 70,000 cells per mL of plasma; the equivalent of a total of 35,000 cells in 500μL of plasma. (Figure 2c), demonstrating the depletion of background host reads. The relative distribution of species was preserved at all concentrations, suggesting low-depth sequencing allows for detection of microbial species in a high-human-background matrix.

**Figure 2.**
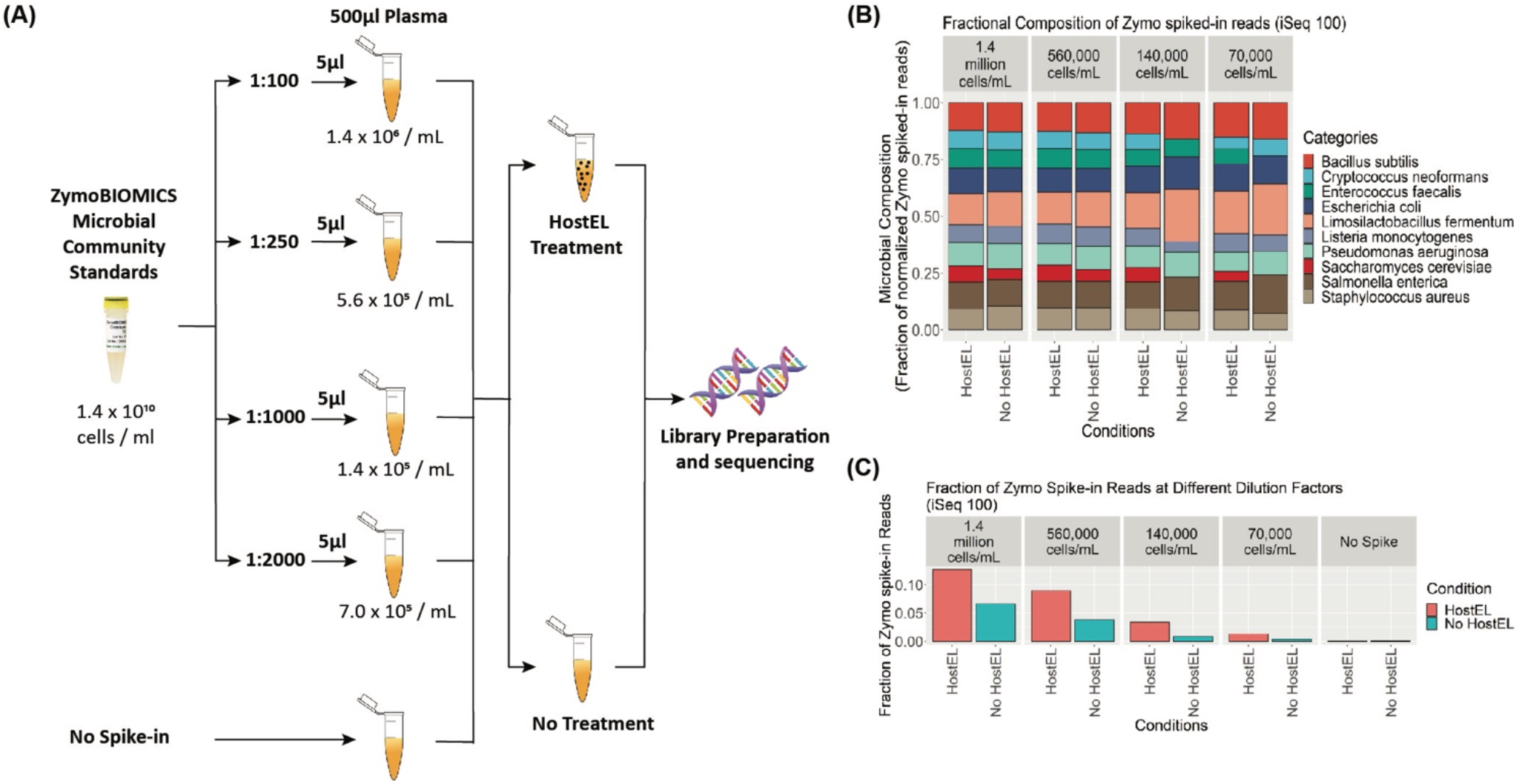
(A) Serial dilution of physiologically relevant levels of the ZymoBIOMICS Microbial Community Standard containing 8 bacteria and 2 yeast (Supplementary Table 1) to the estimated total microbial cells / ml concentration in plasma is done before treatment with or without HostEL followed by TNA extraction and AmpRE library preparation. 5 samples are batched and sequenced on the iSeq 100. (B) Fractional sequencing reads of microbial spike-ins across different dilution factor of HostEL-treated and control samples mapping to the 10 organisms. The relative ratio of the organisms was preserved in the HostEL treated samples at all dilutions, while the control samples lost some organism reads at higher dilutions. (C) Fraction of reads that mapped to spiked-in microbes at different dilutions with or without HostEL treatment. HostEL-treated samples significantly increase the percentage of reads across all different dilution factors as compared to the non-treated samples. Detailed percentages of the reads is shown in Supplementary Table 2.

### HostEL enhances sensitivity and specificity of low depth sequencing

To establish analytical sensitivity and specificity of our approach, we prepared and sequenced 11 ‘blank’ plasma samples and 4 samples each at the above-mentioned dilutions with and without HostEL treatment. Using a stringency metric that is based on the minimum number of read counts to call the presence of an organism to reduce the chance of false positive, we evaluated the sensitivity and specificity of the platform at detecting all 10 spiked-in species as a whole (10 species per sample). With HostEL treatment, we achieved a sensitivity of 96% and a specificity of 96% while the values for the control were 82% and 97% respectively (Figure 3a). Further breakdown of the sensitivity at different dilutions demonstrate the effect HostEL treatment has on the sensitivity of detection, with the lowest concentration registering 88% against 63% of the control sample (Figure 3b). In conclusion, the mNGS workflow shown here enables the detection of bacterial and fungal pathogens with low-depth sequencing.

**Figure 3.**
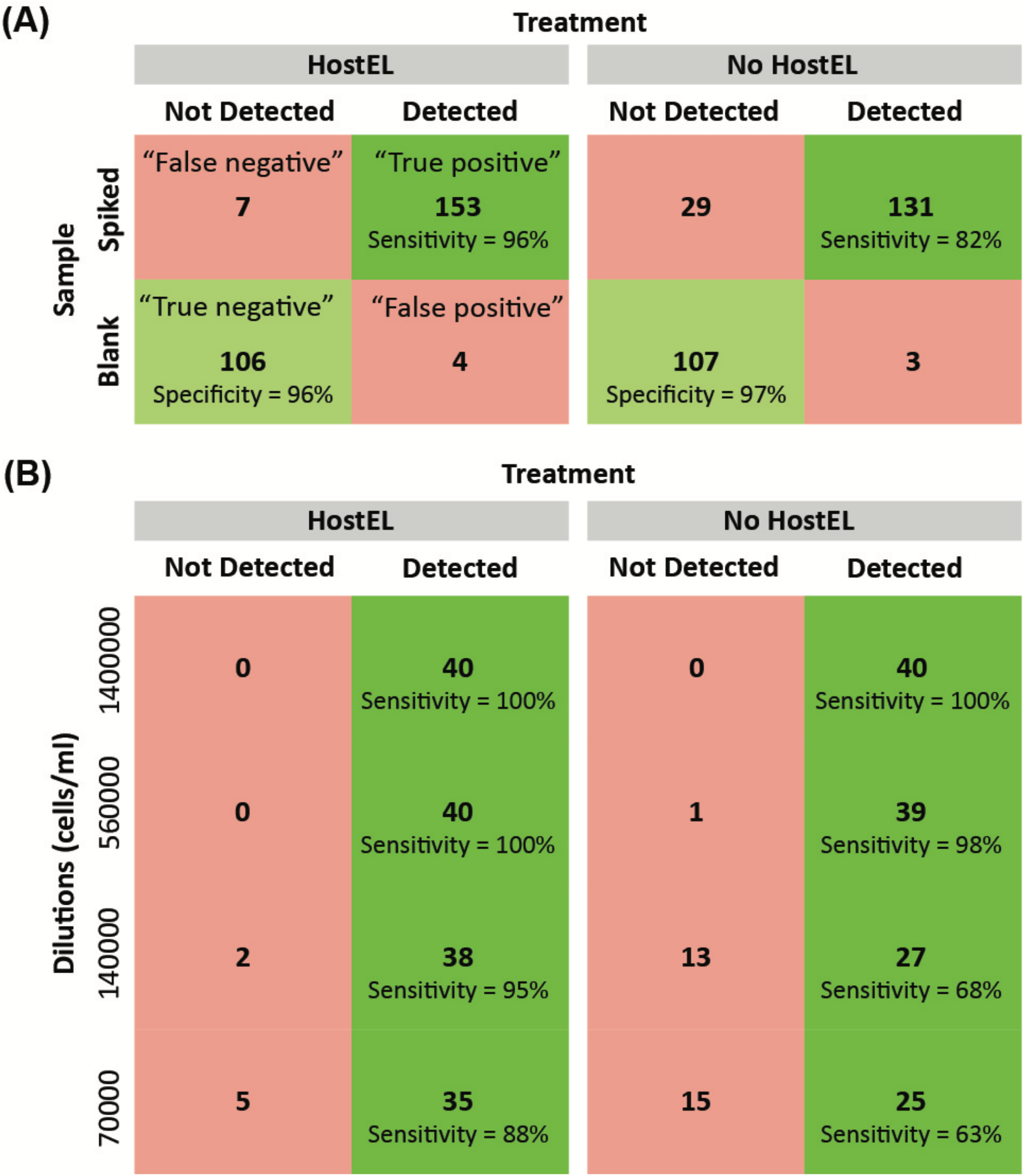
11 blank plasma and 4 repeats of each dilution of the microbial standards were prepared and sequenced on the iSeq 100 with a high stringency cutoff to prevent false positives. Based on the 10 organisms spiked into the plasma, a diagnostic truth table was constructed (A) with and without HostEL treatment and further broken down into (B) detailed sensitivity at different dilutions.

### Concordance with banked patient plasma samples is >90%

Clinical validation of our mNGS workflow was conducted on 42 patient plasma samples collected in compliance to IRB guidelines. These samples have previously been clinically diagnosed with nucleic acid tests to be positive (34 samples) or negative (8 samples) for pathogen infections (Table 1). To understand the correlation between sequencing workflow and the Ct values, and the LOD of the sequencing platforms, qPCR was conducted on 8 clinically molecularly diagnosed Hepatitis B samples with targeted primers (Supplementary Table 3). Correlation plots were plotted with Ct values against the sequencing reads and linear correlation was observed (Figure 4a). As we are targeting 100-500k reads per sample, we can only confidently detect microbial sequences with an abundance of 1 in every 10k reads. For samples with Ct∼30, the relative abundance is about 1 in 10k reads, thus we have high confidence of our sensitivity up to a corresponding Ct value of 30 and below (Supplementary Table 4). When qPCR validation was also done on all sequence-negative clinical samples as quality controls (Table 1) as well as some sequencing-positive samples as positive controls. qPCR was unable to detect any pathogen in 3 out of 13 of these samples, while the rest had a Ct≥33. The high Ct values for these samples may be due to the following reasons: (i) pathogens may be present in a low level; (ii) sample integrity may have been impacted or samples had degraded due to long term storage.

**Table 1.** List of pathogens detected from clinical diagnosis, iSeq 100 and MiniSeq. Green: Clinically diagnosed pathogen was detected on iSeq 100/MiniSeq; Red: Clinically diagnosed pathogen was not detected on iSeq 100/MiniSeq; Blue: Ct value <30; Yellow: Ct value >30 or negative (undetectable).

| Sample | Clinical Diagnosis | iSeq 100 Detection | iSeq 100 Count | MiniSeq Detection | MiniSeq Count | qPCR Ct |
| --- | --- | --- | --- | --- | --- | --- |
| B012 | Hepatitis C Virus | Hepatitis C Virus | 29 | Hepatitis C Virus | 72 | 23.8 |
| B013 | Hepatitis C Virus | Hepatitis C Virus | 32 | Hepatitis C Virus | 66 | 27.2 |
| B014 | HIV-1 | HIV-1 | 10 | HIV-1 | 26 | - |
| B015 | HIV-1 | HIV-1 | 2 | HIV-1 | 3 | - |
| B016 | Hepatitis B Virus | Hepatitis B virus | 4238 | Hepatitis B virus | 11394 | 17.9 |
| B017 | Hepatitis B Virus | Hepatitis B virus | 495 | Hepatitis B virus | 1226 | 24.4 |
| B018 | Hepatitis E Virus | - | 0 | - | 0 | 34.3 |
| B019 | Hepatitis E Virus | - | 0 | - | 0 | 37.1 |
| B020 | Cytomegalovirus | Cytomegalovirus | 9 | Cytomegalovirus | 10 | - |
| B021 | Cytomegalovirus | Cytomegalovirus | 22 | Cytomegalovirus | 100 | - |
| B022 | Varicella Zoster | - | 0 | - | 0 | 33.0 |
| B023 | BK virus | - | 0 | - | 0 | 32.0 |
| B024 | BK virus | - | 0 | - | 0 | 33.0 |
| B025 | Epstein-Barr virus | Epstein-Barr virus | 13 | Epstein-Barr virus | 2 | - |
| B040 | Dengue type 4 | - | 0 | - | 0 | Neg |
| B041 | Dengue type 2 | Dengue virus | 2 | Dengue virus | 3 | - |
| B042 | Dengue type 1 | - | 0 | - | 0 | Neg |
| B043 | Dengue type 3 | Dengue virus | 7 | Dengue virus | 18 | - |
| B045 | Hepatitis B Virus | Hepatitis B virus | 7427 | Hepatitis B virus | 23336 | 22.1 |
| B046 | Hepatitis B Virus | - | 0 | - | 0 | Neg |
| B047 | Hepatitis B Virus | - | 0 | - | 0 | 43.8 |
| B048 | Hepatitis B Virus | Hepatitis B virus | 2 | Hepatitis B virus | 2 | 38.6 |
| B049 | Hepatitis B Virus | Hepatitis B virus | 118 | Hepatitis B virus | 318 | 29.5 |
| B050 | Hepatitis B Virus | - | 0 | - | 0 | Neg |
| B051 | Hepatitis C Virus | Hepatitis C Virus | 17 | Hepatitis C Virus | 128 | 23.3 |
| B052 | Hepatitis C Virus | Hepatitis C Virus | 6 | Hepatitis C Virus | 8 | 26.7 |
| B053 | Hepatitis C Virus | Hepatitis C Virus | 2 | Hepatitis C Virus | 20 | 24.8 |
| B054 | Hepatitis C Virus | - | 0 | - | 0 | 35.0 |
| B056 | Hepatitis C Virus | Hepatitis C Virus | 4 | - | 0 | 32.9 |
| B057 | Hepatitis C Virus | - | 0 | - | 0 | 35.9 |
| B058 | Hepatitis C Virus | - | 0 | - | 0 | 34.1 |
| B059 | Hepatitis C Virus | Hepatitis C Virus | 7 | Hepatitis C Virus | 2 | 28.8 |
| B060 | Hepatitis C Virus | Hepatitis C Virus | 8 | Hepatitis C Virus | 2 | 27.9 |
| B061 | Hepatitis C Virus | - | 0 | - | 0 | 30.1 |
| B068 | No Infection | - | 0 | - | 0 | - |
| B069 | No Infection | - | 0 | - | 0 | - |
| B070 | No Infection | - | 0 | - | 0 | - |
| B071 | No Infection | - | 0 | - | 0 | - |
| B072 | No Infection | - | 0 | - | 0 | - |
| B073 | No Infection | - | 0 | - | 0 | - |
| B074 | No Infection | - | 0 | - | 0 | - |
| B075 | No Infection | - | 0 | - | 0 | - |

**Figure 4.**
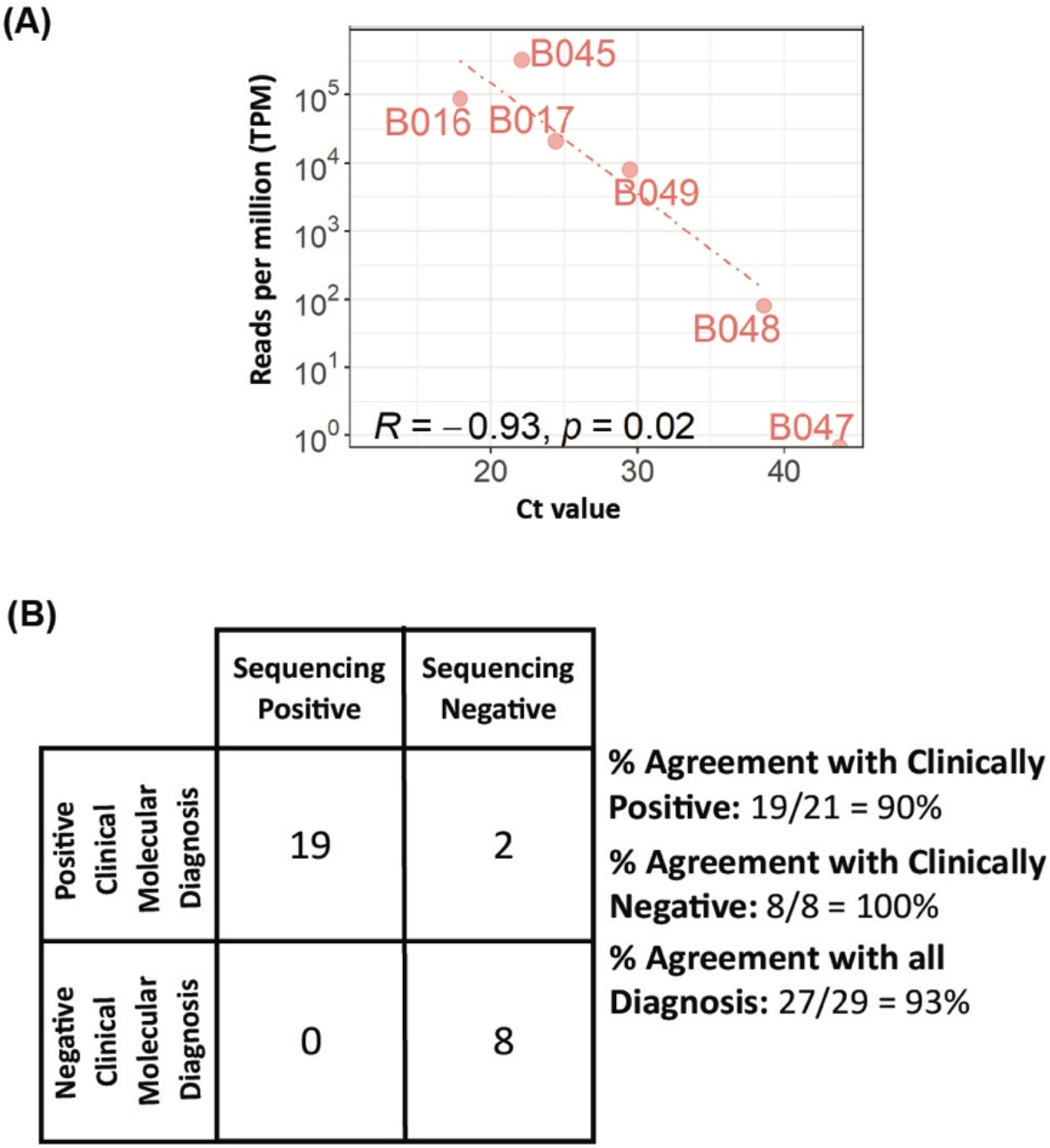
Clinical validation of HostEL and AmpRE workflow. 42 banked patient plasma samples of infected patients or control plasma was obtained from NUS and 500μl of the plasma was put through the workflow just as before. The only difference is that the extraction kit used which resulted in a much larger elution volume. The same 7μl was processed by AmpRE and 5 samples were batched and sequenced on iSeq 100. (A) Correlation between sequencing reads and qPCR results for Hepatitis B-positive samples (Supplementary Table 4). We conducted a qPCR on 8 diagnosed Hepatitis B samples to check the correlation between sequencing reads and pathogen loads. The Ct values were plotted against the sequencing reads. Sequencing reads of both sequencing platforms correlate linearly with the Ct values. For samples with Ct≥33, both sequencing platforms can only pick up less than 10 per million reads, or have stochastic dropout of these pathogens (Table 1). (B) Analysis of clinical validation of HostEL and AmpRE workflow on plasma samples. Our cohort showed concordance results of 100% agreement between the workflow for all the positive clinical molecular diagnosis plasma samples below Ct 33. To ensure there is no false positive, we conducted the workflow on 8 negative clinical molecular diagnosed samples and obtained 100% agreement.

We further compare the percentage agreement of our workflow with the positive clinical molecular diagnosis data and after excluding the 13 samples that were deemed to be low quality (Ct≥33 or undetectable) from the statistics (Figure 4b). We obtained 93% agreement with the clinical results. To ensure that there is no false positive in our workflow, 8 control plasma samples (B068-B075) clinical negative for pathogen were also analyzed with the workflow (Table 1). Our workflow shows 93% agreement on iSeq 100 on both positive and negative samples with Ct<33 (Figure 4b). In rare cases with Ct≥33 where sample quality is low (B048), pathogen detection is still possible although with a lower sensitivity (Table 1).

### Both DNA and RNA viruses can be detected if Ct is less than 33

Both DNA viruses (Hepatitis B, CMV, EBV) and RNA viruses (Hepatitis C, Dengue, HIV) were detected from the clinical samples (Table 1), suggesting that the one-pot AmpRE library preparation kit enables the detection of both DNA and RNA pathogens. Unlike the analytical validation when the extracted TNA was eluted in 30μl, the clinical samples were eluted in 60μl of water based on clinical SOP. With only 7μl used as input into the library preparation, we were effectively sampling 58μl of plasma (500μl x 7μl / 60μl) to detect the pathogen. With a more aggressive concentration strategy during nucleic acid extraction, the sensitivity will likely increase.

### Rapid turnaround can be achieved with different sequencing platfors

Our workflow was tested on the iSeq 100 2×150 sequencing platform due to its ease of use, which would be a big factor in clinical deployment of a metagenomic sequencing solution. However, if implemented in its entirety, the whole workflow will deliver the results to the infectious disease specialist in 27 hours (Figure 1c). In theory, with a sample submission cutoff at 11am and samples loading at 5pm, the report will be out at 2pm the next day with our workflow. If the sequencing time was reduced to 13 hours, then it is possible to deliver results by 8am the next morning forward rounds. To simulate such a workflow, we trimmed the reads to 2 × 100 paired end to see if that affected the sensitivity and specificity of such a low-depth approach to metagenomic sequencing (‘iSeq-trimmed’). In addition, we evaluated the MiniSeq, which can deliver a single end 100 nucleotide read in 5 hours, potentially shortening the entire workflow to 13 hours, albeit with the trade-off of greater user involvement in setting up the MiniSeq run. To compare the low-depth sequencing, the read count was kept at 1 million reads per sample (MiniSeq: 20 samples per sequencing run; iSeq100: 5 samples per sequencing run).

The analytical validation using 4 (iSeq-trimmed) or 1 sample (MiniSeq) at each dilution level with negative controls obtained comparable sensitivity with the 19 hour protocol for both the trimmed and MiniSeq run (Figure 5a). The MiniSeq, in particular, resulted in a higher sensitivity without HostEL treatment than the iSeq 100 albeit with a much smaller sample size. Although the sample size was small, it suggests the specificity is comparable for all 3 protocols (Supplementary Table 2). Thus, we can conclude that reducing the run time with shorter protocols seems to result in no loss of sensitivity.

**Figure 5.**
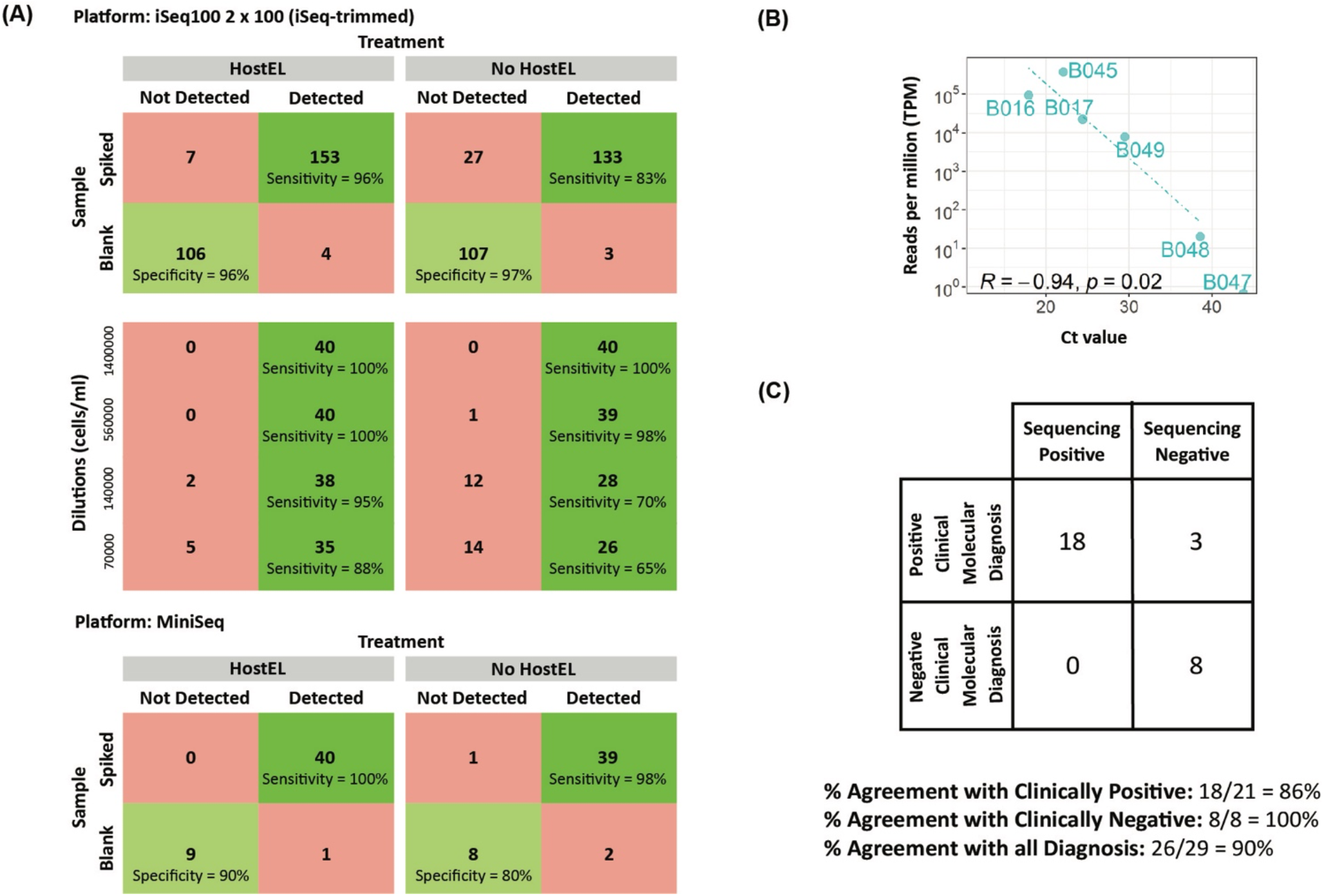
Clinical options for turnaround time. We computationally trimmed the reads of the iSeq 100 to 2 × 100 to simulate a 13 hour run time and also ran the same libraries batched with 20 samples per run on the MiniSeq. (A) Sensitivity and specificity performance on the spiked-in samples for the trimmed iSeq 100 and MiniSeq runs were similar to the full iSeq 100 run. Breakdown of the different dilution levels for iSeq-trimmed is also included. (B) Correlation between sequencing reads and qPCR results for Hepatitis B-positive samples (Supplementary Table 4) (C) MiniSeq diagnostic truth table for samples with Ct<33.

Correlation plots of Ct values against the sequencing reads were also plotted for the miniSeq and linear correlation was also observed (Figure 5b, Supplementary Table 4). With the patient samples and a Ct 33 cutoff, MiniSeq performed equivalently to the iSeq 100 run (Figure 5c). Our results suggest that the HostEL and AmpRE workflow is compatible with all three sequencing setup and can deliver rapid unbiased metagenomic sequencing results with shallow sequencing.

## Discussion

Metagenomic next-generation sequencing (mNGS) has been used extensively in the detection of infectious diseases and is found to have certain advantages over conventional methods. It is an unbiased approach that can detect both known and unknown pathogens. Huge amount of effort has been made in key steps of mNGS, such as human background depletion, non-human sequence enrichment, library preparation, etc., to improve the diagnosis efficiency (10,35–42). Nevertheless, challenges such as cost and simplicity of mNGS method still exist(3). In this study, we addressed these challenges by establishing a workflow that uses HostEL and AmpRE kits for human background depletion and library preparation.

There are a few challenges involving in human background depletion. Firstly, the soluble nucleases are hard to remove from the reactions, and second, it is hard to fine tune the preferential lysis. We developed the strategy, HostEL, for preferential lysis of human cells followed by enzymatic digestion of human background using enzyme-immobilized magnetic beads, leaving intact pathogens for total nucleic acids (TNA) extraction and sequencing. As only a single step of magnetic beads removal is needed to remove the enzymes without the need for enzyme deactivation, it can be applied to biofluids of as low as 500μL. This strategy allows easy integration into any clinical system as it is a simpler and shorter process. We validated HostEL in our analytical validation using ZymoBIOMICS Microbial Community Standard (43), consisting of 8 bacteria and 2 yeasts, as a spike-in control to mimic the physiological infection condition. As illustrated in our study, HostEL is able to deplete human nucleic acid without compromising on the composition of the pathogens. At 1.4 million cells/mL of plasma, HostEL is able to increase non-human reads by ∼4-fold. When the microbial contribution was diluted to 70,000 cells/mL of plasma, a similar level to the physiological level of 10^5^ copies/mL as reported in Darton et al., 2009 (30), the composition of the pathogens remained consistent. This appears to be superior to other depletion methodologies available in terms of maintaining relative composition (44). In a separate study, non-human fraction of the sequencing reads in mNGS was found to be as low as less than 0.1% of the total reads (45). Hence with the use of HostEL, we are able to save cost on mNGS as the same amount of data is obtainable from less sequencing data. There is a question on the limit of detection and sensitivity of this approach as opposed to more sensitive qPCR, targeted sequencing approaches or even higher depth unbiased metagenomic sequencing, especially with the Ct33 detection cutoff for the banked clinical samples. While health economics calculations are not beyond the scope of this article, the availability of a first-line diagnostic for febrile neutropenia or fever of unknown origins that can detect an active infection of viruses, bacteria or fungi at a comparatively low price point can save the patient weeks of diagnostic workup and provide timely targeted therapeutic intervention. This has the potential to reduce hospitalization duration and can be beneficial for antibiotic stewardship by only targeting the right organisms, potentially at the expense of missing residual infections that are not detectable in the blood.

Typical strategy for sequencing library construction for DNA and RNA is to have two separate library preparations. The AmpRE kit is to combine both library preparations into a single pot, which reduces the cost since the same reagents are used. As the kit is fast (under 4 hours) and can be used for input as low as 0.7ng of TNA, we are able to obtain adequate signal from 500μL of biofluids after treatment with HostEL. Sequencing of AmpRE generated libraries can be done on demand in small batches on iSeq 100, allowing rapid detection of both DNA and RNA viruses, bacteria and fungus if sufficient samples are obtained daily. At 5 samples per run, we believe the sample load in a tertiary hospital should enable daily sequencing runs with the corresponding rapid speed of result delivery. Currently, AmpRE is optimized to produce libraries of certain sizes that are better for short-read sequencing. However, the strategy can be tuned for other sequencing platforms. Unlike most library preparation kits, AmpRE is also a pure additive process with minimal clean-up steps, thus reducing the time and effort needed. Nonetheless, there is risk of having background amplicons in the samples, and hence our workflow is optimized to prevent such occurrence.

In our clinical validation cohort of 42 plasma samples, which consist of DNA and RNA viruses as well as bacteria (Table 1), HostEL and AmpRE workflows are able to achieve high agreement with the clinical molecular diagnosis data. We performed the sequencing on two separate sequencing platforms, iSeq 100 and MiniSeq, as well as a simulated short iSeq 100 run and achieved concordance results of 90% agreement with the positive clinical molecular diagnosis samples. Our workflow also showed 100% agreement with the negative clinical molecular diagnosis samples, ruling out the high possibility of having false positive result. We further demonstrated that sequencing reads scale linearly with the pathogen load by conducting qPCR on clinically diagnosed Hepatitis B samples. Correlation plots of the Ct value against sequencing reads show that sequencing reads increase linearly with the decreasing Ct values, but the limit of detection of our sequencing platforms dropped when Ct>30. Our initial results of our workflow are promising, however limited to small diversity of pathogens and biofluid due to logistical limitation and timeline. We intend to extend our workflow to larger diversity of pathogens with different specimen types. We acknowledge that there are limitations to our workflow as it is not applicable to parasite infections as parasites will not survive under selective lysis assay.

We also validated this workflow on iSeq 100 and MiniSeq to demonstrate the portability of the assay between different Illumina platforms at different sequencing depths and different run times. While sequencing workflows can theoretically be performed in under 24 hours in ideal conditions, there are real-world limitations such as operational hours of diagnostic laboratories, requirement for batching to minimize sequencing costs, analytical pipeline timing is frequently omitted from the calculations and cost considerations (3). To enable unbiased metagenomic sequencing to be a first-line diagnostic option, there is a need to deliver cost-competitive solutions that fit clinical laboratory setup. In the 3 workflows presented, the relatively short sample preparation time (6 hours) coupled with competitive sequencing costs (Figure 1a), the option of different workflows and rapid sample analysis (2 hours), enable hospitals to offer unbiased metagenomic sequencing as a diagnostic tool with next-day turnaround routinely. The option of the simplified iSeq 100 workflow can work well with labs with less skilled technicians at the cost of longer turnaround while the MiniSeq offers speed at the cost of setup complexity. All 3 protocols have similar detection capabilities and perform well with clinical samples. Thus, we believe the workflow presented in this publication can potentially benefit the clinical community.

In conclusion, we established a mNGS workflow using HostEL and AmpRE, which considerably reduces time, cost and effort, by effectively depleting human background and amplifying both DNA and RNA into sequencing library in a single reaction, as illustrated in our analytical and clinical validations. We foresee to explore fungal infection and also infections in a diversity of biofluids, and anticipate to scale up the number of clinical samples in our validation to further support the efficiency of our strategies in different clinical settings.

## Data Availability

The analytical validation sequencing dataset for this study can be found in the figshare repository 10.6084/m9.figshare.21694184. The clinical validation dataset contains patient genomic data and is available for download upon request pending ethics approval for each individual request.

https://doi.org/10.6084/m9.figshare.21694184

## Author Contributions

WK, SH, SY conceived and designed the study. GY and CKL was involved in selecting the samples used for banked patient samples. SIP, WK and SY performed the sequencing experiments other than HostEL treatment and TNA extraction of the banked patient samples, which was performed by CKL and THMC. LL and SIP performed the qPCR on the banked samples. KWK, WK, SY, WYTL, CC drafted the manuscript.

## Acknowledgments

The authors would like to thank the patients from NUHS for contributing their samples, NUHS for collecting and processing the plasma samples, and Illumina, Inc. for providing access to the MiniSeq system for this study.

## Supplementary Material

### Supplementary Figures

**Supplementary Figure 1.**
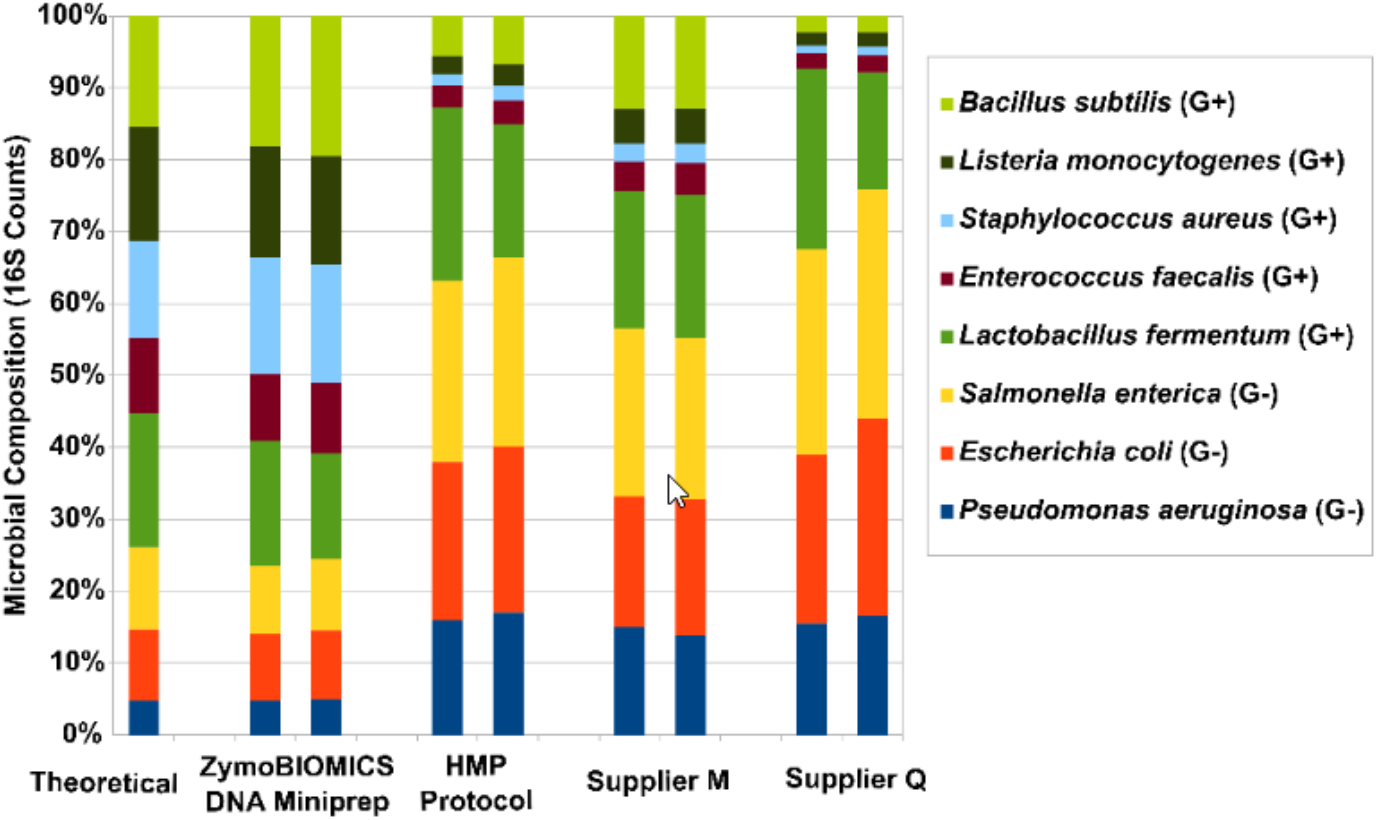
Zymobiomics theoretical spike-in composition based on 16S rRNA amplicon sequencing (1).

### Supplementary Tables

**Supplementary Table 1.** ZymoBIOMICS Microbial Community Standard (Zymo Research, Cat no. D6300) consist of 8 bacterial strains (3 Gram-negative and 5 Gram-positive) and 2 tough-to-lyse yeasts at a concentration of 1.4 × 10^10^ cells/mL (1).

| Species | Theoretical Composition (%) |  |  |  |  | Gram Stain |
| --- | --- | --- | --- | --- | --- | --- |
|  | Genomic DNA | 16S Only | 16S & 18S | Genome Copy | Cell Number |  |
| *Pseudomonas aeruginosa* | 12 | 4.2 | 3.6 | 6.1 | 6.1 | - |
| *Escherichia coli* | 12 | 10.1 | 8.9 | 8.5 | 8.5 | - |
| *Salmonella enterica* | 12 | 10.4 | 9.1 | 8.7 | 8.8 | - |
| *Lactobacillus fermentum* | 12 | 18.4 | 16.1 | 21.6 | 21.9 | + |
| *Enterococcus faecalis* | 12 | 9.9 | 8.7 | 14.6 | 14.6 | + |
| *Staphylococcus aureus* | 12 | 15.5 | 13.6 | 15.2 | 15.3 | + |
| *Listeria monocytogenes* | 12 | 14.1 | 12.4 | 13.9 | 13.9 | + |
| *Bacillus subtilis* | 12 | 17.4 | 15.3 | 10.3 | 10.3 | + |
| *Saccharomyces cerevisiae* | 2 | NA | 9.3 | 0.57 | 0.29 | Yeast |
| *Cryptococcus neoformans* | 2 | NA | 3.3 | 0.37 | 0.18 | Yeast |

**Supplementary Table 2.**
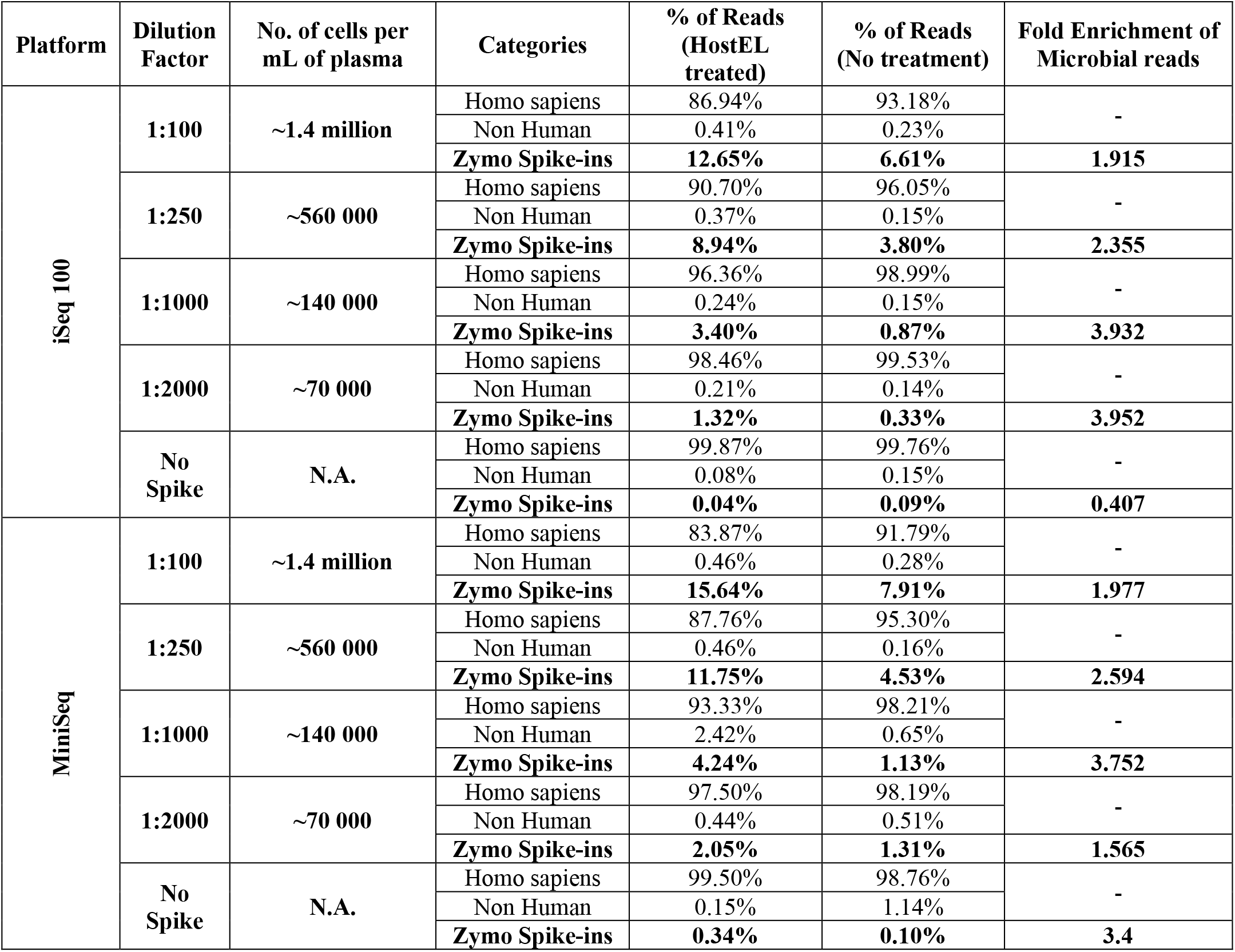
Detailed breakdown of percentage of reads of HostEL treated vs no treatment enrichment on iSeq 100 and MiniSeq sequencing platforms. HostEL can enrich the percentage of reads by ∼2 fold in plasma with high microbial concentrations (1.4 million cells/mL) and ∼4 fold with low microbial concentration (140,000 cells/mL and 70,000 cells/mL) on iSeq 100 sequencing platform while enriching at least 1.5 fold on MiniSeq sequencing platform.

**Supplementary Table 3.**
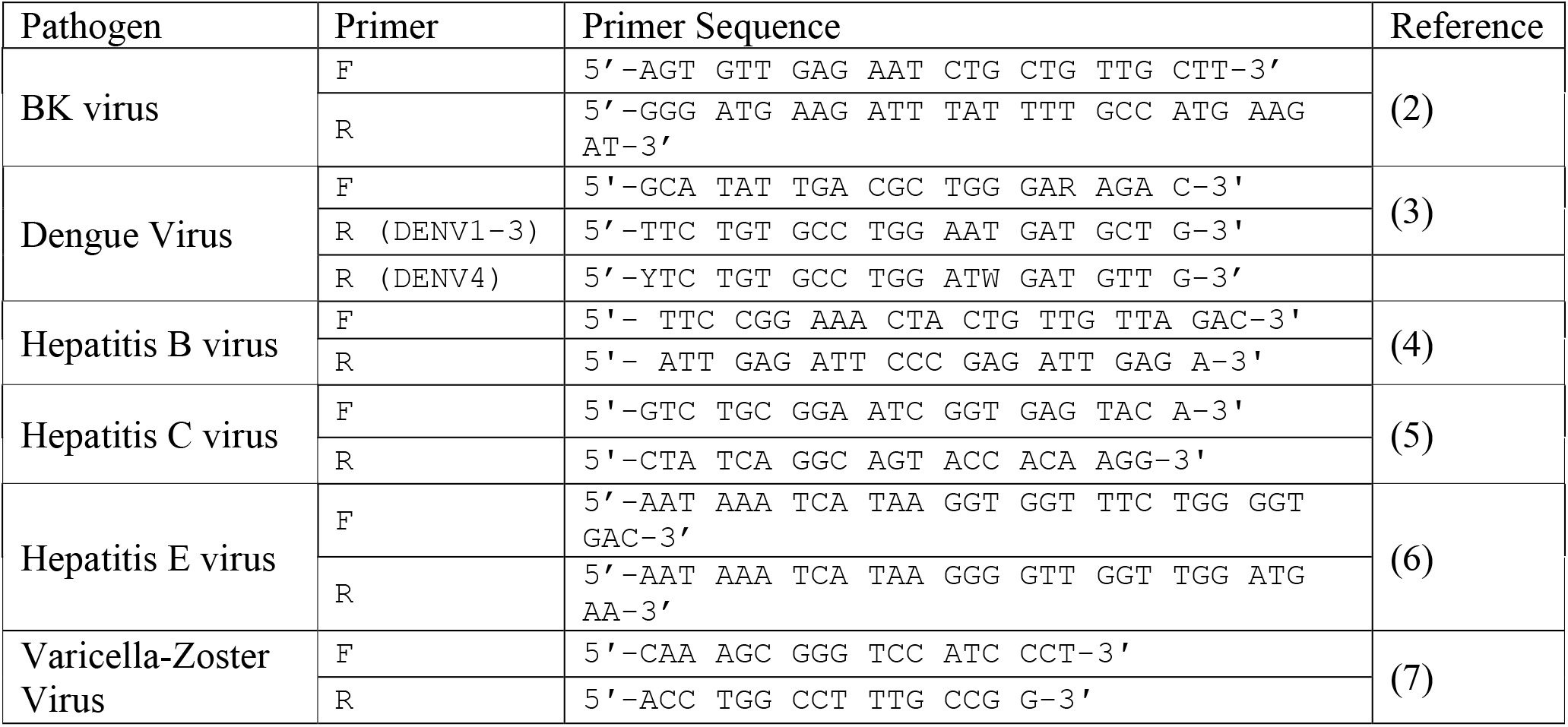
List of targeted primers used in qPCR validation of clinical samples.

**Supplementary Table 4.** qPCR and iSeq 100/MiniSeq sequencing results of Hepatitis B plasma samples

| Samples | qPCR Ct Value | Frac Reads | Raw Reads Counts | Platform | Clinical Diagnosis |
| --- | --- | --- | --- | --- | --- |
| B047 | 43.8 | 0.00E+00 | 0 | iSeq 100 | Hepatitis B virus |
| B046 | UD | 0.00E+00 | 0 |  |  |
| B050 | UD | 0.00E+00 | 0 |  |  |
| B048 | 38.6 | 8.00E-05 | 2 |  |  |
| B049 | 29.5 | 8.05E-03 | 118 |  |  |
| B017 | 24.4 | 2.11E-02 | 495 |  |  |
| B016 | 17.9 | 8.64E-02 | 4238 |  |  |
| B045 | 22.1 | 3.32E-01 | 7427 |  |  |
| B047 | 43.8 | 0.00E+00 | 0 | MiniSeq |  |
| B046 | UD | 0.00E+00 | 0 |  |  |
| B050 | UD | 0.00E+00 | 0 |  |  |
| B048 | 38.6 | 2.00E-05 | 2 |  |  |
| B049 | 29.5 | 7.80E-03 | 318 |  |  |
| B017 | 24.4 | 0.02203 | 1226 |  |  |
| B016 | 17.9 | 0.09309 | 11394 |  |  |
| B045 | 22.1 | 3.77E-01 | 23336 |  |  |

